# Temporal trends in preterm birth rate over the last 30 years in Sweden: a population-based study

**DOI:** 10.64898/2026.02.24.26346962

**Authors:** B. Han, H. Sundelin, K. Ytterberg, J. Juodakis, P. Nyeboe, A. Rosengren, U. Strömberg, M. Norman, T. Svanvik, P. Solé-Navais, B. Jacobsson

## Abstract

**Objectives:** To determine temporal trends in the rates of preterm birth and its sub-types in Sweden and assess the contribution of known-risk factors.

**Design:** A population-based register study.

**Setting:** Sweden.

**Participants (Instead of patients or subjects):** 3,264,146 pregnancies registered in the Swedish Medical Birth Registry with information on pregnancy duration and onset of labour (1991 – 2021).

**Main outcome measures:** The primary outcomes were the overall, spontaneous and iatrogenic preterm birth rates between 1991 - 2021, stratified on singleton and multiple births, as well as for extremely preterm (<28 weeks, <196 days), very preterm (28-31 weeks, 196 - 224 days), moderately preterm (32 - 33 weeks, 224 - 238 days), and late preterm (34 - 36 weeks, 238 - 259 days) births. Using logistic regression models, we investigated whether maternal age at conception, use of artificial reproductive technologies, smoking, parity, and maternal continent of birth were associated with the observed trends.

**Results:** The overall preterm birth rate was stable between 1991 - 2005 at 5.50% (95% CI: 5.37%, 5.63% in 1991) but decreased thereafter to 4.78% (95% CI: 4.66%, 4.91%) in 2021, a finding confined to spontaneous preterm births. The largest decline was observed in late preterm births, from 3.92% (95% CI: 3.80%, 4.05%) in 2005 to 3.52% (95% CI: 3.41%, 3.63%) in 2021. Moderately preterm birth also declined (0.70%, 95% CI: 0.65%, 0.76% in 2005 to 0.53%; 95% CI: 0.49%, 0.58% in 2021), whereas very-extremely preterm birth did not. Decreased spontaneous preterm birth rates were observed in women born in European, Asian and African countries, with largest decline observed in the latter (rate in 1991 = 2.65%, 95% CI: 1.74%, 3.86%; rate in 2021 = 1.72%, 95% CI: 1.42%, 2.07%). Adjusting for maternal and obstetric risk factors didn’t alter these associations.

**Conclusions:** While rates of preterm birth have been stable or increased globally, they have decreased in Sweden from 2006 - 2021, despite the lack of any nation-wide preventive strategy during this period. Understanding the reasons for this decline will provide useful strategies to make the decline a rule, rather than an exception.

## Introduction

Preterm birth, defined as a birth before 37 weeks of gestation, is the leading cause of perinatal mortality worldwide and a major contributor to the global burden of disease [1]. Each year, approximately 13.4 million babies are born preterm, accounting for around 10% of all births [2].

Despite improved living conditions, better antenatal care [3] and a decrease in known risk factors, such as smoking during pregnancy [4], global rates of preterm birth have remained high for the past decades [5–7]. Changes in maternal characteristics, including increasing maternal age [8–11] and the growing use of assisted reproductive technologies [12–14] are potential contributors to such stalling rates in high-income countries. However, the extent to which these and other factors explain temporal trends in preterm birth is not fully understood. Examining long-term patterns within individual countries can therefore help clarify how demographic and obstetric changes may influence preterm birth rates.

Sweden offers an exemplary setting to examine such trends. The country maintains systematically collected, high-quality data through the Swedish Medical Birth Register [15,16], including information on gestational age, birth subtypes, maternal demographics, and lifestyle factors such as nicotine use.

In this population-based register study, we investigated secular trends in overall, spontaneous, and iatrogenic preterm birth in Sweden between 1991 and 2021, stratified by multiples/singleton and detailed gestational-age categories. By describing long-term patterns prior to the introduction of national preventive guidelines [17], this study aims to provide insights into the roles that demographic, obstetric and reproductive factors may have in the secular trends in preterm birth.

## Materials and Methods

### Ethical approval

This project was approved by the Swedish Ethical Review Authority (Dnr. 2024-08620-02), allowing the use of pseudonymized individual-level data from the Swedish Medical Birth Register. Because of the nature of the data, informed consent was waived.

### Sample selection

This study used data from the Swedish Medical Birth Register, which contains almost all births in Sweden since 1973, with detailed information such as labour onset, complications during pregnancy, neonatal period, birth, reproductive history and demographic information [15,16]. In this study, all pregnancies that resulted in a live birth between 154 and 308 days of gestation (22 and 44 weeks), from January 1, 1991, to December 31, 2021 was included. Prior to 1991, information regarding the onset of labour was not registered. Pregnancies with missing gestational duration, stillbirths, mothers without pseudonymized identifier, mother-child pairs with duplicated pseudonymized identifier, and outlier pregnancies based on the correlation of birth weight and gestational age (using generalized additive models for location scale and shape (GAMLSS) [18]) were excluded (Figure 1). Importantly, twins and other multiples were considered as a single pregnancy (i.e., counting pregnancies instead of births). After filtering, we analysed data from a total of 3,210,294 pregnancies in 1,736,391 mothers.

**Figure 1.**
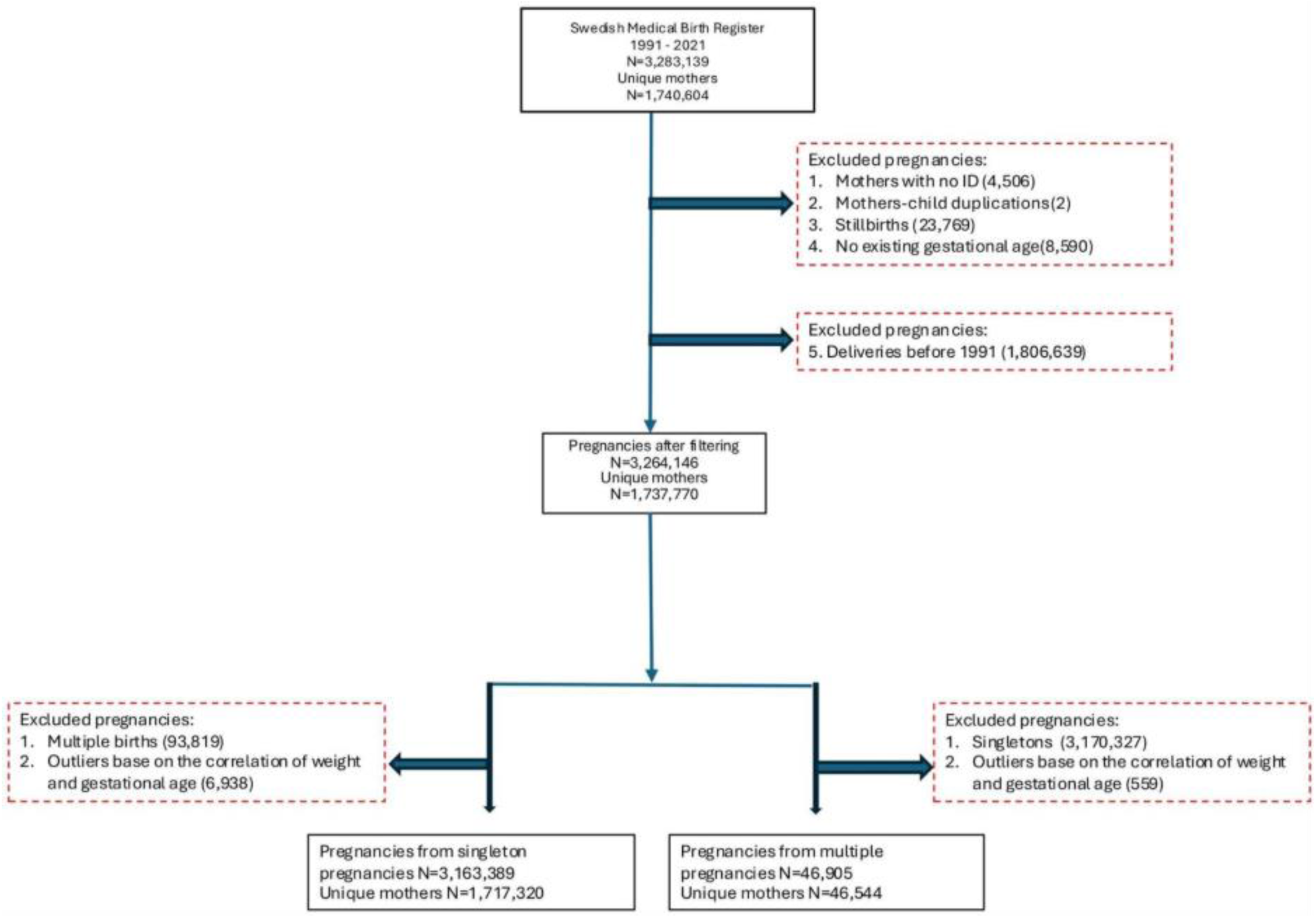
Flowchart of the stepwise data cleaning process of the Swedish Medical Birth Register.

### Variable definitions

In this study, the “best estimate” of gestational duration according to the Swedish Medical Birth Register was used^16^, which is based on either ultrasound (85% of the records) or the date of the last menstrual period. We also present sensitivity analyses using “LMP-based estimate” of gestational duration, which is calculated as the difference in days between birth date and the last menstrual period. Pregnancies ending before 259 days (37 weeks) were considered as preterm births, regardless of how labour was initiated or whether the pregnancy carried singletons or multiples. As mentioned above, we counted the number of pregnancies ending in a preterm birth, and not the number of births per se; a preterm twin pregnancy contributes with one additional preterm birth, and not two. The preterm birth rate was defined as the number of preterm births divided by the total number of births (i.e., pregnancies). At the same time, we also studied the rate of spontaneous preterm birth and iatrogenic preterm birth, as well as those for singleton and multiple births. Spontaneous preterm birth included all births <259 days (<37 completed weeks) after spontaneous onset of contractions with or without prelabor rupture of membranes. Iatrogenic preterm births were births initiated by either induction of labour or caesarean section (elective or emergency).

Finally, preterm birth were split into five categories accordingly to gestational duration: extremely preterm (<28 weeks, <196 days), very preterm (28 - 31 weeks, 196 - 224 days), moderately preterm (32 - 33 weeks, 224 - 238 days) and late preterm (34 - 36 weeks, 238 - 259 days).

The main outcomes were the total rate of preterm birth, spontaneous preterm birth, and iatrogenic preterm birth. For all outcomes, the denominator was the total number of births, the number of births in singletons, or the number of births in multiples.

Additional data on maternal age at conception, usage of artificial reproductive technologies, smoking, parity, and maternal origin (continent of birth) were also extracted from the Swedish Medical Birth Register.

### Statistical analysis

To estimate the rate of change in preterm birth rates across years, we fit a linear model to the annual rates of preterm birth obtained. Based on an observation that the rates were different in two halves of the study period, the linear model was used separately for the years 1991 - 2005 and 2006 - 2021, but note that no statistical test was applied to this split. This was performed for both singletons and multiples.

For estimating the preterm birth risk associated with maternal country of birth, we fit a linear model for the spontaneous preterm birth rate in women of European, Asian and African origin for the periods 1991-2021.

Finally, we estimated the associations between year and preterm birth risk by using the years 1991 - 1995 as reference and applying logistic regression models on individual level data. We display the odds ratio for giving birth preterm in unadjusted models, after adjusting for maternal age at conception and parity, further adjusting for smoking, use of artificial reproductive technologies, and maternal continent of origin.

All analyses were undertaken using the R language (version 4.4.2) and the code used for the study is available at https://github.com/PerinatalLab/time-trend-PTD.

## Results

### Study population characteristics

For 1991 - 2021 there were a total of 3,264,146 pregnancies recorded in the Swedish Medical Birth Register. After predefined exclusions, our final study population comprised 3,210,294 pregnancies (3,163,389 singletons and 46,905 multiples, **Figure 1**). Maternal age at conception increased steadily during the period 1991 - 2021 (**Supplementary Figure 1**), with the mean maternal age being 27.38 (95% CI: 27.37, 27.39) in 1991 and 30.35 (95% CI: 30.33, 30.36) in 2021. The smoking rate during pregnancy decreased steadily (**Supplementary Figure 2**) during the period 1991 - 2021, from 24.25% (95% CI: 24.00%, 24.50%) in 1991 to 3.25% (95% CI: 3.15%, 3.36%) in 2021, while the use of artificial reproductive technologies increased from 3.70% (95% CI: 3.58%, 3.82%) in 1995 (registering of the use of artificial reproductive technologies started in 1995) to 12.33% (95% CI: 12.13%, 12.52%) in 2021. The rate of nulliparous women remained relatively stable from 1991 to 2021 (**Supplementary Figure 2**). The rate of mothers with Asian background increased from 2.97% (95% CI: 2.87%, 3.07%) to 12.92% (95% CI: 12.73%, 13.12%). The rate of mothers with African background increased from 0.81% (95% CI: 0.76%, 0.87%) to 5.69% (95% CI: 5.55%, 5.83%) (**Supplementary Figure 3**). The rate of multiple pregnancies (**Supplementary Figure 4**) appears to have increased from 1.25% (95% CI: 1.18%, 1.31%) in 1991 to 1.66% (95% CI: 1.58%, 1.74%) 2003, with a subsequent decrease and stagnation at ∼1.41% from 2004 and onwards. This sudden decline and stagnation coincided with the establishment of a Swedish national recommendation for single embryo transfer in artificial reproductive technology. Finally, the rate of iatrogenic births (i.e., including inductions and caesarean sections) has increased steadily, from 22.00% (95% CI: 21.77%, 22.23%) in 1991 to 35.84% (95% CI: 35.55%, 36.12%) in 2021 (Supplementary Figure 5).

### Preterm birth rates decreased after 2006 in Sweden

For each calendar year, the proportions (with 95% confidence intervals, 95% CI) of preterm birth in all pregnancies, in singletons and in multiples were estimated. Overall preterm birth rate was stable during the period 1991-2005 and steadily decreased after that. Preterm birth rate was 5.50% (95% CI: 5.37%, 5.63%) in 1991 and decreased to 4.78% (95% CI: 4.66%, 4.91%) in 2021. During 1991 - 2005, the total preterm birth rate remained stable (beta: 0.002%; p-value: 0.803) and thereafter decreased by 0.00047 per year (p-value: 2.967e-11; **Figure 2A and Supplementary Table 1**). Preterm birth rates for singletons have similar trends compared to the overall preterm birth rate, because singleton pregnancies contribute most of the total number of births each year (**Figure 2B** and **Supplementary Table 2**). Singleton preterm birth rates were 5.01% (95% CI: 4.88%, 5.13%) and 4.27% (95% CI: 4.15%, 4.39%) in 1991 and 2021, respectively. Between 1991 - 2005, the singleton preterm birth rates remained stable (beta: - 0.003%; p-value: 0.739) and thereafter decreased by 0.00047 (p-value: 2.967e-11) per year. A similar trend in multiple pregnancies (**Figure 2C** and **Supplementary Table 3**) was observed, with the multiples preterm birth rate being stable for the years 1991 - 2005 (beta: -0.048%; p-value = 0.401) and decreasing thereafter, from 43.09% (95% CI: 40.54%, 45.67%) in 2006 to 42.13% (95% CI: 39.61%, 44.68%) in 2021, a decrease of 0.00147 per year (p-value: 0.063).

**Figure 2.**
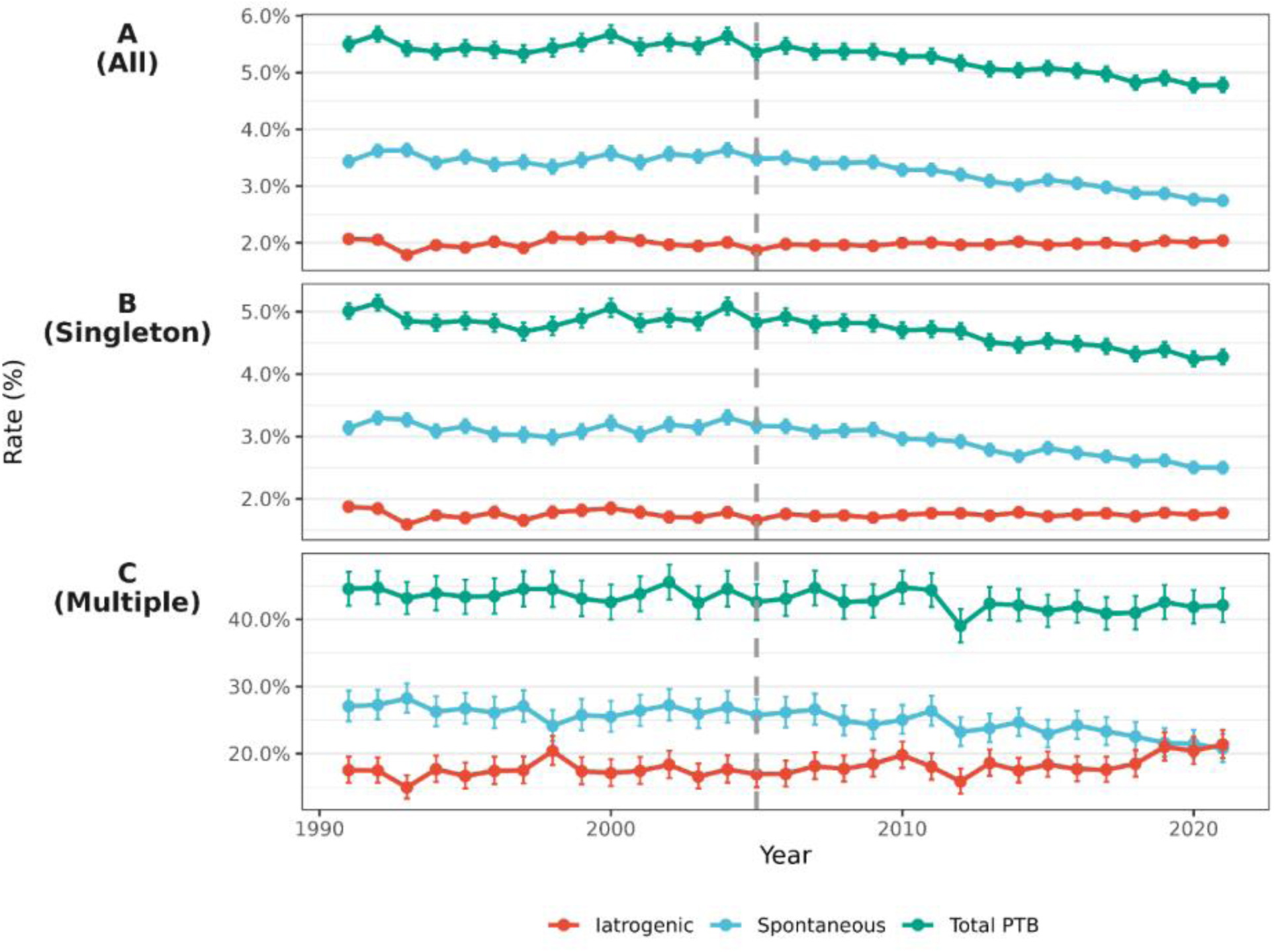
Preterm birth rate for A) all, B) singleton and C) multiple pregnancies in Sweden between 1991 - 2021. Rates of preterm birth were calculated as the number of preterm births over the total number of all the births, singletons and multiples that year. The error bars represent the 95% CI and the vertical dotted, grey line represents year 2005.

We attribute the decline in preterm birth rate to a reduction in the rate of spontaneous preterm births, whereas the iatrogenic preterm birth rate remained stable during the whole study period (1991 - 2021) for all, singleton and multiple pregnancies, see **Figure 2**.

### The decrease is observed among births occurring from 32 weeks of gestation

To evaluate trends of preterm birth over time in different sub-categories, annual rates of extremely preterm (<28 weeks, <196 days), very preterm (28 - 31 weeks, 196 - 224 days), moderately preterm (32 - 33 weeks, 224 - 238 days) and late preterm (34 - 36 weeks, 238 - 259 days) births were calculated. For each calendar year, the proportion of the different sub-categories of preterm birth was estimated. We observed a decline, to varying degrees, in the rates of preterm birth in all sub-categories of preterm birth, except for the extremely preterm births (**Figure 3)**. The strongest decline was observed in late preterm births (34 - 36 weeks), with a decline from 4.02% (95% CI: 3.91%, 4.13%) in 1991 to 3.52% (95% CI: 3.41%, 3.63%) in 2021, followed by moderately preterm birth (32 - 33 weeks), (0.72%; 95% CI: 0.67%, 0.77% in 1991 to 0.53%; 95% CI: 0.49%, 0.58% in 2021). The decline was weak, and non-significant, for very preterm births (28 - 31 weeks) (0.54%; 95% CI: 0.50%, 0.58% in 1991 to 0.48%; 95% CI: 0.44%, 0.52% in 2021). Finally, a weak, non-significant increase in the rate of extremely preterm births (< 28 weeks), from 0.22% (95% CI: 0.19%, 0.25%) in 1991 to 0.25% (95% CI: 0.22%, 0.28%) in 2021 was observed. Similar trends were observed even if only singletons or multiple pregnancies were considered (**Supplementary Figure 6 and Supplementary Figure 7**).

**Figure 3.**
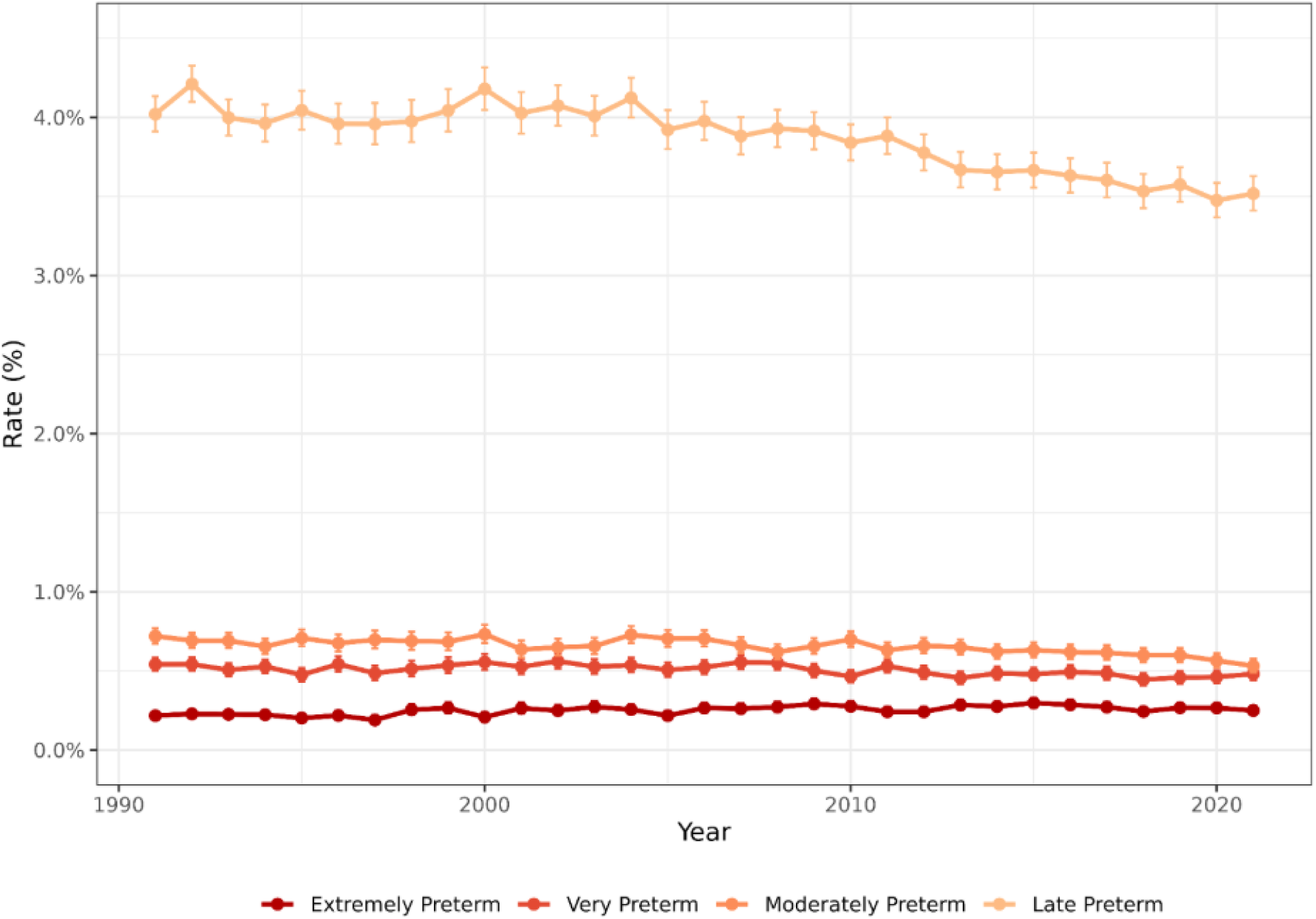
Rates of extremely preterm (<28 weeks), very preterm (28 - 31 weeks), moderately preterm (32 – 33 weeks), and late preterm (34 - 36 weeks) births in Sweden between 1991 - 2021. Rates of preterm birth were unadjusted, calculated as the number of preterm births by the total number of births in that year. The error bars represent the 95% CI.

In singleton pregnancies, the rate of late preterm (34 - 36weeks), showed a significant decline from 3.71% (95% CI: 3.61%, 3.82%) in 1991 to 3.19% (95% CI: 3.08% 3.29%) in 2021 (**Supplementary Figure 6**), while the decline in the rate of moderately preterm births (32 - 33 weeks), was weaker, ranging from 0.63% (95% CI: 0.59%, 0.68%) in 1991 to 0.45% (95% CI: 0.41%, 0.49%) in 2021. The rate of very or extremely preterm births (28 – 31 or < 28 weeks), remained stable throughout the study period.

In multiple pregnancies (**Supplementary Figure 7**), the rate of late preterm birth (34 - 36 weeks) declined from 28.51% (95% CI: 26.24%, 30.87%) in 1991 to 28.00% (95% CI: 25.73%, 30.35%) in 2021, while that of moderately preterm births (32 - 33 weeks) showed a slight-significant decline, ranging from 7.73% (95% CI: 6.43%, 9.20%) in 1991 to 6.43% (95% CI: 5.24%, 7.80%) in 2021 and that of very preterm births (28 - 31 weeks) declined from 5.86% (95% CI: 4.73%, 7.17%) in 1991 to 5.29% (95% CI: 4.21%, 6.55%) in 2021. Similarly to the group of singleton pregnancies, the rates of extremely preterm births (< 28 weeks) remained stable throughout the study period, ranging from 2.47% (95% CI: 1.74%, 3.38%) in 1991 to 2.41% (95% CI: 1.69%, 3.32%) in 2021.

### Declines in preterm birth rates were observed in mothers from European, Asian and African origin

Maternal country of birth has previously been associated with preterm birth risk [19]. Now, we examined the temporal trends in preterm birth rates in women from European, Asian and African origin (i.e., maternal country of birth in one of these continents). The preterm birth rate used in this study is defined as the rate of spontaneous preterm births among all births in the corresponding background population. A decreasing trend in spontaneous preterm birth rate was observed in all three groups (**Figure 4**). The strongest decrease in preterm birth rate was observed in mothers of African origin, in whom rates decreased from 2.65% (95% CI: 1.74%, 3.86%) in 1991 to 1.72% (95% CI: 1.42%, 2.07%) in 2021, a decrease of 0.04% per year (p-value =1.932e-07) (**Supplementary Table 4**). A similar decreasing trend was noticed in mothers of Asian origin, in whom spontaneous preterm birth decreased from 3.66% (95% CI: 3.07%, 4.33%) in 1991 to 2.81% (95% CI: 2.55%, 3.10%) in 2021, resulting in a total decrease of 0.032% per year (p-value=6.40e-05). The spontaneous preterm birth rate in mothers of European origin also decreased throughout the studied time-period, but more modestly, ranging from 3.42% (95% CI: 3.31%, 3.53%) in 1991 to 2.80% (95% CI: 2.69%, 2.91%) in 2021, a decrease of 0.022% per year (p-value =7.959e-09). Given the relatively few pregnancies of mothers born in Oceania, North America and South America during the years 1991 - 2021, we excluded these mothers from the analysis.

**Figure 4.**
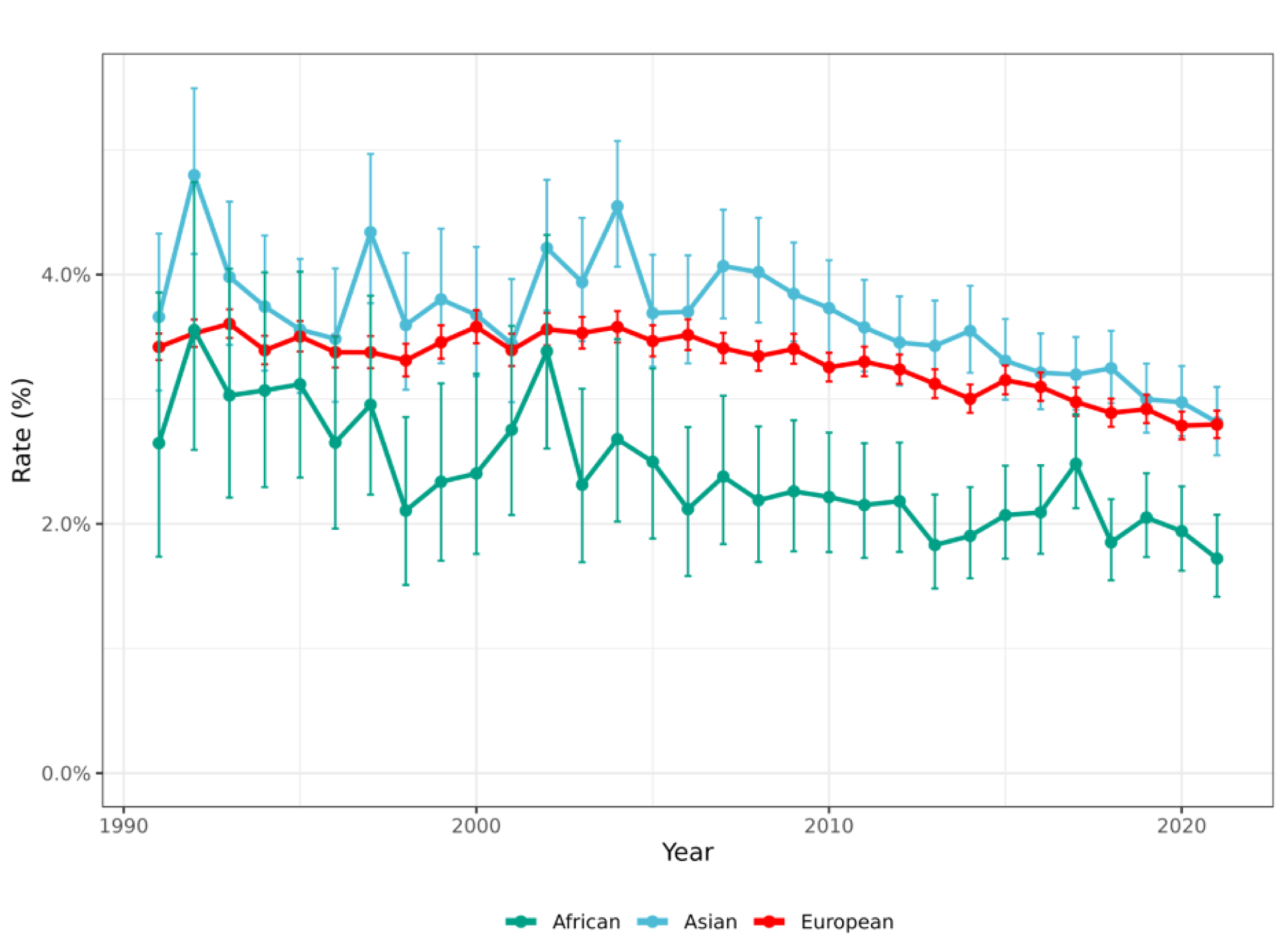
Spontaneous preterm birth rates in mothers born in Africa, Asia and Europe giving birth in Sweden during the time-period 1991 – 2021. Error bars indicate 95% CI.

### The decline in preterm birth rates is robust to adjustments for traditional risk factors

The relations between the prevalence of risk factors and the observed decline in the preterm birth rates between 2005 and 2021 were studied. Logistic regression models were used to estimate the odds ratios of spontaneous preterm birth in different birth year groups (1991 - 2021), without adjusting, and further adjusting for maternal age, parity, smoking prior to pregnancy, maternal continent of birth, and use of assisted reproductive technology. In 3,026,900 singleton births (86,839 spontaneous preterm births), we observed that adjusting for maternal age, parity, use of artificial reproductive technologies and maternal continent of birth had no effects on the estimated decline in preterm birth rates between 2006 - 2021 **(Figure 5)**. Further adjusting for maternal smoking prior to pregnancy slightly attenuated the decline but did not account for the total decrease in preterm birth rates reported above.

**Figure 5.**
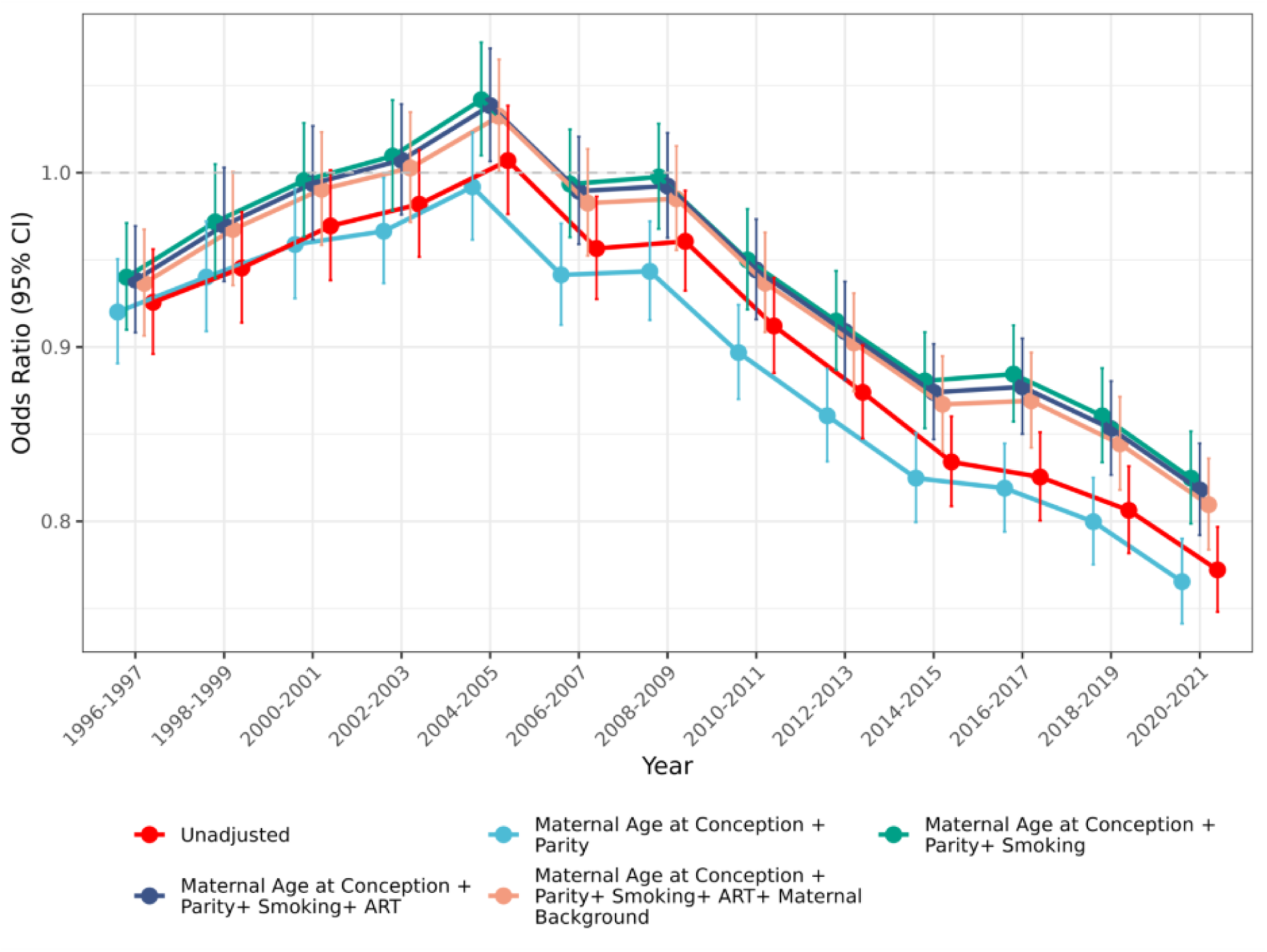
Odds ratio for preterm birth with vertical error bars indicating 95% CI for giving birth in one of the following years after 1996 compared to the years 1991-1995 (reference). Analyses are shown unadjusted and after adjusting for maternal age at conception, parity, smoking before pregnancy, use of artificial reproductive technologies, and maternal continental origin.

### Spontaneous preterm birth rates decreased in Sweden also if gestational duration was dated using last menstrual period

Changes in gestational duration dating practices within the study period could have contributed the observed time-trends in preterm birth rates. Therefore, we decided to conduct sensitivity analyses by estimating the time-trends of preterm birth rates using gestational duration estimated using last menstrual period. For each calendar year, we estimated the proportions (with 95% CI) of preterm birth by the method of last menstrual period in singletons and assessed. The singleton preterm birth rate was 4.49% (95% CI: 4.37%, 4.62%) in 1991 and 4.53% (95% CI: 4.41%, 4.66%) in 2021. The preterm birth rate remained stable during the whole study period (1991 - 2005: beta: 0.0065%; p-vale: 0.327; 2006 - 2021: beta: -0.0008% per year; p-value: 0.852; **Figure 6 and Supplementary Table 5**). Singleton spontaneous preterm birth rates were 2.80% (95% CI: 2.70%, 2.91%) and 2.56% (95% CI: 2.47%, 2.66%) between 1991 and 2021, respectively. Between 1991 - 2005, the preterm birth rates remained stable (beta: 0.0043%; p-value: 0.468) and thereafter decreased by 0.025% (p-value: 1.988e-06) per year. Singleton iatrogenic preterm birth rates were 1.69% (95% CI: 1.61%, 1.77%) and 1.97% (95% CI: 1.88%, 2.05%) between 1991 and 2021, respectively. Between 1991 - 2005, the preterm birth rates remained stable (beta: 0.0021%; p-value: 0.674) and thereafter increased by 0.0264% (p-value: 2.346e-07) per year.

**Figure 6.**
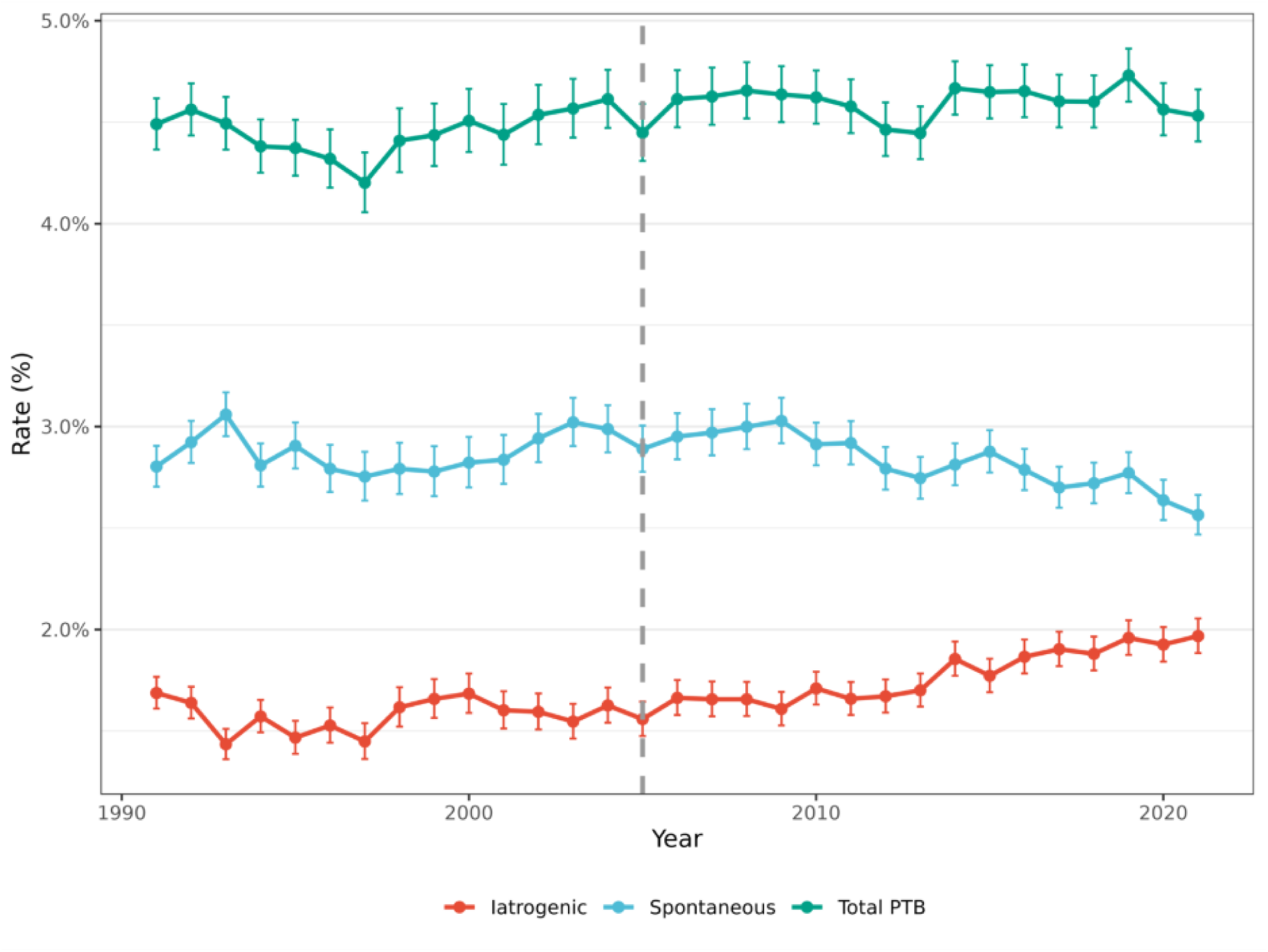
Preterm birth rate for singleton pregnancies in Sweden between 1991 – 2021, with gestational duration estimated using last menstrual period. Rates of preterm birth were calculated as the number of preterm births over the total number of singletons that year. The error bars represent the 95% CI and the vertical dotted, grey line represents year 2005.

A decreasing trend in spontaneous preterm birth rate by last menstrual period was observed in women from European, Asian and African origin (**Supplementary Figure 8**). Different with the results by best estimate, the strongest decrease in preterm birth rate was observed in mothers of Asian origin, in whom rates decreased from 3.77% (95% CI: 2.98%, 4.68%) in 1991 to 2.49% (95% CI: 2.22%, 2.77%) in 2021, a decrease of 0.032% per year (p-value =6.059e-05) (**Supplementary Table 6 and Supplementary Figure 8**).

## Discussion

In this study, we investigated the national trends in preterm birth rates across Sweden from 1991 to 2021. We observed a consistent decline in preterm birth rate in Sweden beginning in 2005 and continuing through 2021, driven primarily by a reduction among births with spontaneous onset, and in those occurring from 32 weeks of gestation. This decline was consistently observed in women born in Europe, Asia and Africa was not explained by temporal changes in recognized risk factors for preterm birth.

In the recent Born Too Soon report, global rates of preterm birth are reported to have remained largely unchanged across all regions around 9.9% [6,7]. Globally, preterm birth rates have shown little evidence of decline over recent decades and have remained largely stable or increased in many countries, despite sustained recognition of preterm birth as a major public health concern [5–7]. Notably, increases in preterm birth rates were reported between 2010 and 2020 in several countries, including the United Kingdom (including Northern Ireland), Ireland, and Iceland [5–7]. We have previously reported a marked increase in preterm birth rates in Sweden following the introduction of routine ultrasound dating of pregnancy in the early 1980s, followed by a prolonged period of relative stability from the mid-1980s through the early 2000s [20–23]. The Netherlands is another country where preterm birth rates have declined recently [24]. Nonetheless, reductions have been driven primarily by decreases in iatrogenic preterm births, particularly at later gestational ages, whereas spontaneous preterm birth has remained stable or increased at earlier gestations.

The predominance of the reduction in spontaneous preterm birth is of particular interest, as spontaneous preterm birth is generally considered less amenable to prevention than medically indicated preterm birth and has shown little decline in many other high-income settings. The observed decrease therefore suggests the influence of broader, population-level changes rather than targeted clinical interventions. Potential contributors may include improvements in maternal health, antenatal care practices, or unmeasured social or environmental factors [25]. Importantly, the decline predates the introduction of Sweden’s first national guidelines for the medical prevention of preterm birth in 2023 [17], underscoring the need to identify and understand the mechanisms underlying this favorable trend.

Previous studies have shown that temporal changes in maternal demographic, behavioral, and clinical risk factors explain only a limited proportion of secular trends in preterm birth rates [26–29]. Against this background, given shifts in population risk factors over time, it is plausible that opposing trends may partly offset one another and influence overall patterns in preterm birth.

However, in analyses examining the association between these shifts and the observed decline in spontaneous preterm birth between 2006 and 2021, we found no evidence that changes in established risk factors explained the reduction in the Swedish dataset. Adjustment for maternal age, parity, use of assisted reproductive technologies, and maternal continent of birth had no effect on the estimated decline, and additional adjustment for smoking prior to pregnancy only slightly attenuated the association without accounting for the overall decrease. Together, these findings indicate that changes in traditional population-level risk factors do not explain the observed decline in preterm birth, suggesting that other, as yet unidentified, determinants are likely contributing to this trend.

An alternative explanation for the observed temporal decline in preterm birth rate might therefore be related to a shift in gestational age estimation methods. The Swedish Medical Birth Register is a high-quality, population-based register with near-complete coverage and detailed information on multiple approaches to estimating gestational duration, including last menstrual period and ultrasound-based dating and an algorithm for best estimate of gestation duration [30,31]. By the late 2000s, manual gestational dating procedures had largely been eliminated from the registry, tempering observable variation and strengthening the reliability of gestational age estimates^22^. Improvements in ultrasound technology, personnel training, and greater use of first trimester rather than second-trimester ultrasound dating may also introduce temporal variation that complicates the interpretation of secular trends in preterm birth. In the present study, we therefore used different sensitivity analyses to estimate the effect of gestational dating methods. The decline in spontaneous preterm birth rates was observed regardless of the method used to date the pregnancy. Nevertheless, the precise timing and magnitude of the decline are more difficult to quantify, as estimates based on ultrasound and last menstrual period differ somewhat.

A birth cohort effect of the women in the registry has to be considered. While the women giving birth in the early 1990s will have been born from the 1950s to the 1980s, the most recent mothers in the study were born on average more than 30 years later. Their early year experiences will have been markedly different with respect to child and school, health care, diet, and exposure to teenage smoking, which was rampant in the 1960s and 1970s, but may not have been adequately recorded. Still, this exposure was likely only present in Swedish-born women and potentially ruling out birth cohort effects [32]. Trends in passive smoking will also need to be considered in future studies [33], with continuously lower exposure over time. Notably, smoking in restaurants and nightclubs was forbidden in 2005, indicating a substantially lower exposure in young women since.

A major strength of this study is the use of Sweden’s nearly complete national perinatal registry, which captures approximately 96% of all births and includes nearly 3.2 million births between 1991 and 2021 [30]. The high quality and detailed structure of the Medical Birth Register enabled differentiation between spontaneous and iatrogenic onset of birth, singleton and multiple pregnancies, and assessment of several established risk factors for preterm birth, including maternal continent of birth. However, smoking status is self-reported and therefore subject to misclassification, and the extent of missing data may vary across regions and over time [30]. A key limitation of the study is that no combination of measured risk factors could account for the observed decline in preterm birth rates in Sweden. Future studies should therefore consider the role of non-traditional or less well-captured risk factors. Finally, the absence of data on neonatal complications and the timing of ultrasound examinations used for gestational dating limits the ability to fully assess the clinical implications of the observed trends. Whether the observed reduction in spontaneous preterm birth is accompanied by corresponding improvements in short-and long-term infant outcomes remains to be established.

### Conclusion

While preterm birth rates have increased globally, this study demonstrates a sustained decline in preterm birth in Sweden between 2005 and 2021, occurring in the absence of nationwide preventive interventions and without being explained by reductions in the prevalence of established risk factors. Whether other countries represent similar exceptions to prevailing global trends remains to be determined; however, identifying the factors underlying the Swedish decline may offer valuable insights for transforming such reductions from isolated occurrences into a broader, reproducible pattern.

## Funding

Funding was provided by the FORTE, Swedish Research Council for Health, Working Life and Welfare (ALFGBG-1005149), Stockholm, Sweden (2023-00253), the Swedish Society for Medical Research (SG-24-0105-B) and the Swedish Research Council, Stockholm, Sweden (2019-01004, 2023-02735 and 2024-02502).

## Transparency

Language in this manuscript has been edited with the assistance of an artificial intelligence (AI) tool for language clarity, grammar, and stylistic improvement. The authors take full responsibility for the content of the text.

## Data Availability

All data produced in the present study are available upon reasonable request to the authors

## Acknowledgements

None.

## Author contributions

BH was involved in conceptualization, data curation, formal analysis, methodology, data visualisation and writing the original draft of the manuscript. HS and KY were involved in data curation and methodology. JJ and PS were involved in conceptualization, methodology, and software and supervision. BJ was involved in conceptualization, funding acquisition, methodology, project administration, resources and supervision. All authors were involved in writing, reviewing and editing the manuscript. The corresponding author attests that all listed authors meet authorship criteria and that no others meeting the criteria have been omitted. BH is the guarantor.

## Supplementary Tables

**Supplementary Table 1.** Changes in preterm birth rates in Sweden in 1991-2005 and 2006-2021.

| Onset of labour | Time period | Estimate | p-value | Std. Error |
| --- | --- | --- | --- | --- |
| All births | 1991-2005 | 0.00002 | 0.803 | 6.949e-05 |
| All births | 2006-2021 | -0.00047 | 2.97e-11 | 2.525e-05 |
| Spontaneous | 1991-2005 | 0.00002 | 0.688 | 5.985e-05 |
| Spontaneous | 2006-2021 | -0.00050 | 7.611e-12 | 2.443e-05 |
| Iatrogenic | 1991-2005 | -0.000007 | 0.904 | 5.605e-05 |
| Iatrogenic | 2006-2021 | 0.000033 | 0.033 | 1.382e-05 |
| Linear model over the summarized data of preterm birth rate between 1991 and 2021. Model was unadjusted. |  |  |  |  |

**Supplementary Table 2.** Changes in preterm birth rates in singleton pregnancies.

| Onset of labour | Time period | Estimate | p-value | Std. Error |
| --- | --- | --- | --- | --- |
| All births | 1991-2005 | -0.00003 | 0.739 | 7.883e-05 |
| All births | 2006-2021 | -0.00047 | 2.967e-11 | 2.525e-05 |
| Spontaneous | 1991-2005 | 0.000006 | 0.929 | 6.280e-05 |
| Spontaneous | 2006-2021 | -0.00050 | 7.611e-12 | 2.443e-05 |
| Iatrogenic | 1991-2005 | -0.00003 | 0.527 | 5.007e-05 |
| Iatrogenic | 2006-2021 | 0.00003 | 0.033 | 1.382e-05 |
| Linear model over the summarized data of preterm birth rate between 1991 and 2021. Model was unadjusted. |  |  |  |  |

**Supplementary Table 3.** Changes in preterm birth rates in multiple pregnancies.

| Onset of labour | Time period | Estimate | p-value | Std. Error |
| --- | --- | --- | --- | --- |
| All births | 1991-2005 | -0.00048 | 0.401 | 0.00056 |
| All births | 2006-2021 | -0.00147 | 0.063 | 0.00073 |
| Spontaneous | 1991-2005 | -0.00081 | 0.173 | 0.00056 |
| Spontaneous | 2006-2021 | -0.00326 | 4.591e-06 | 0.00045 |
| Iatrogenic | 1991-2005 | 0.00032 | 0.646 | 0.00069 |
| Iatrogenic | 2006-2021 | 0.00179 | 0.020 | 0.00068 |
| Linear model over the summarized data of preterm birth rate between 1990 and 2021. Model was unadjusted. |  |  |  |  |

**Supplementary Table 4.** Preterm birth changes for mothers from European, Asian and African throughout the whole study period.

| **Background** | **Estimate** | **p-value** | **Std. Error** |
| --- | --- | --- | --- |
| Africa | -0.00040 | 1.932e-07 | 5.943e-05 |
| Asia | -0.00032 | 6.40e-05 | 6.861e-05 |
| Europe | -0.00022 | 7.959e-09 | 2.754e-05 |
| Estimates were obtained from a linear regression model over summarized data of preterm birth rate in singletons between years 1991-2021 |  |  |  |

**Supplementary Table 5.** Changes in preterm birth rates in singleton pregnancies by last menstrual period.

| **Onset of labour** | **Time period** | **Estimate** | **p-value** | **Std. Error** |
| --- | --- | --- | --- | --- |
| All births | 1991-2005 | 0.000065 | 0.327 | 6.353e-05 |
| All births | 2006-2021 | -0.000008 | 0.852 | 4.125e-05 |
| Spontaneous | 1991-2005 | 0.000043 | 0.468 | 5.793e-05 |
| Spontaneous | 2006-2021 | -0.00025 | 1.988e-06 | 3.271e-05 |
| Iatrogenic | 1991-2005 | 0.000021 | 0.674 | 4.981e-05 |
| Iatrogenic | 2006-2021 | 0.000246 | 2.346e-07 | 2.647e-05 |
| Linear model over the summarized data of preterm birth rate between 1991 and 2021. Model was unadjusted. |  |  |  |  |

**Supplementary Table 6.** Preterm birth changes by last menstrual period for mothers from European, Asian and African throughout the whole study period.

| **Background** | **Estimate** | **p-value** | **Std. Error** |
| --- | --- | --- | --- |
| Africa | -0.00030 | 6.393e-05 | 6.411e-05 |
| Asia | -0.00032 | 1.366e-05 | 6.059e-05 |
| Europe | -0.00023 | 6.887e-11 | 2.259e-05 |
| Estimates were obtained from a linear regression model over summarized data of preterm birth rate in singletons between years 1991-2021 |  |  |  |

## Supplementary Figures

**Supplementary Figure 1.**
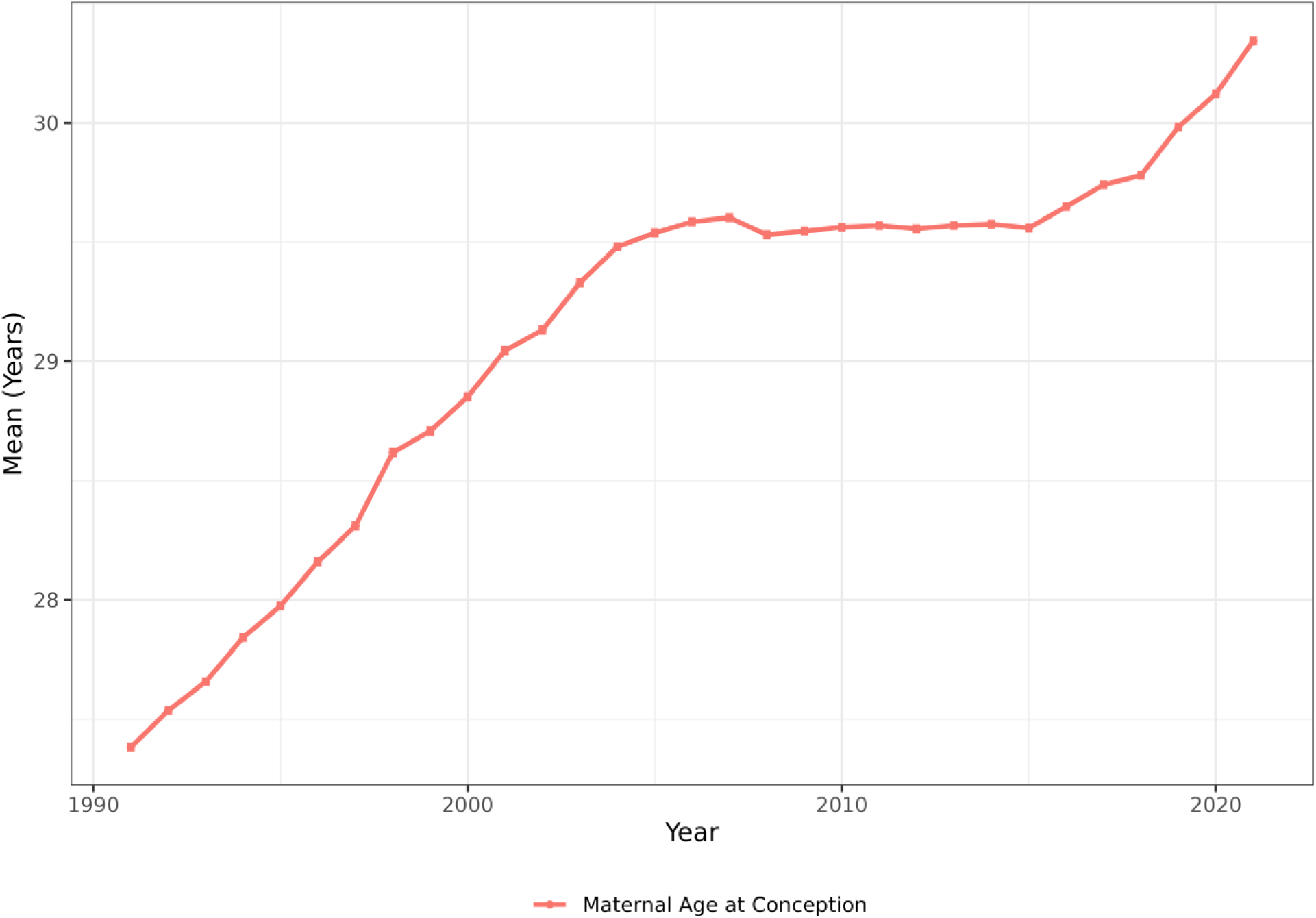
Change in mean maternal age at conception in Sweden from 1991 to 2021. Error bars indicate 95% CI.

**Supplementary Figure 2.**
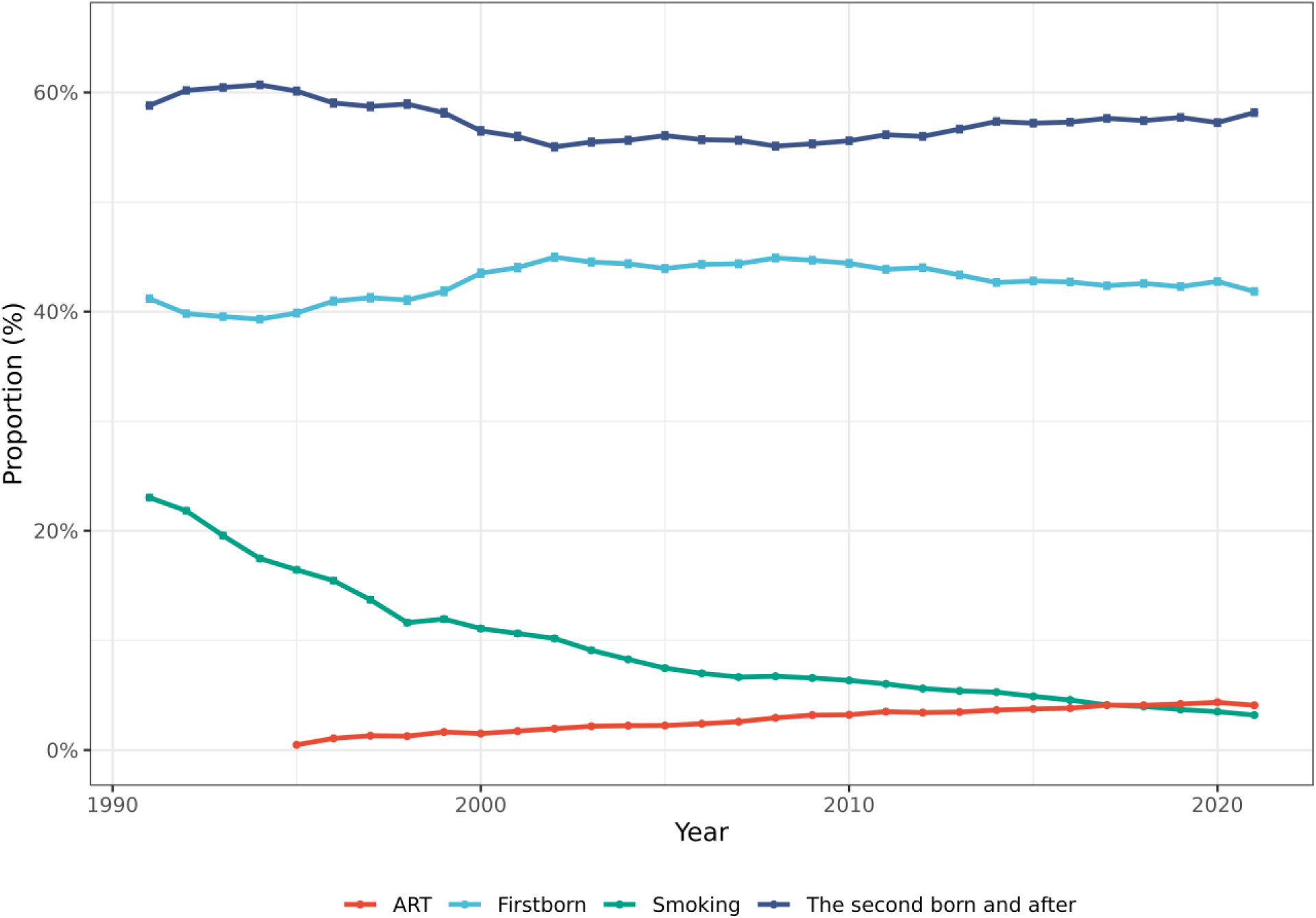
Rates of parity, smoking prior to pregnancy, use of assisted reproductive technologies in Sweden from 1991 to 2021. Error bars indicate 95% CI.

**Supplementary Figure 3.**
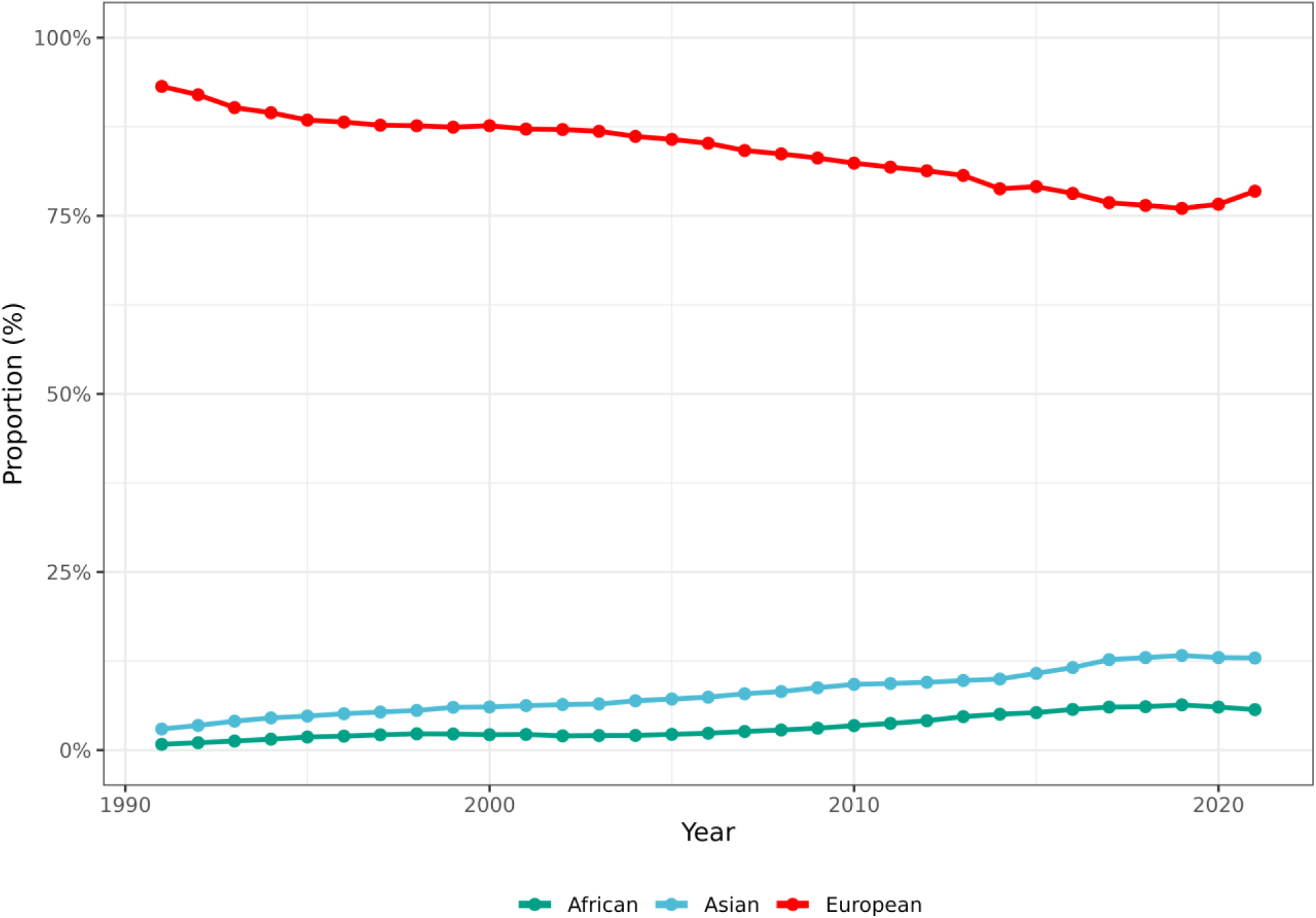
The rates of mothers from different backgrounds. Error bars indicate 95% CI.

**Supplementary Figure 4.**
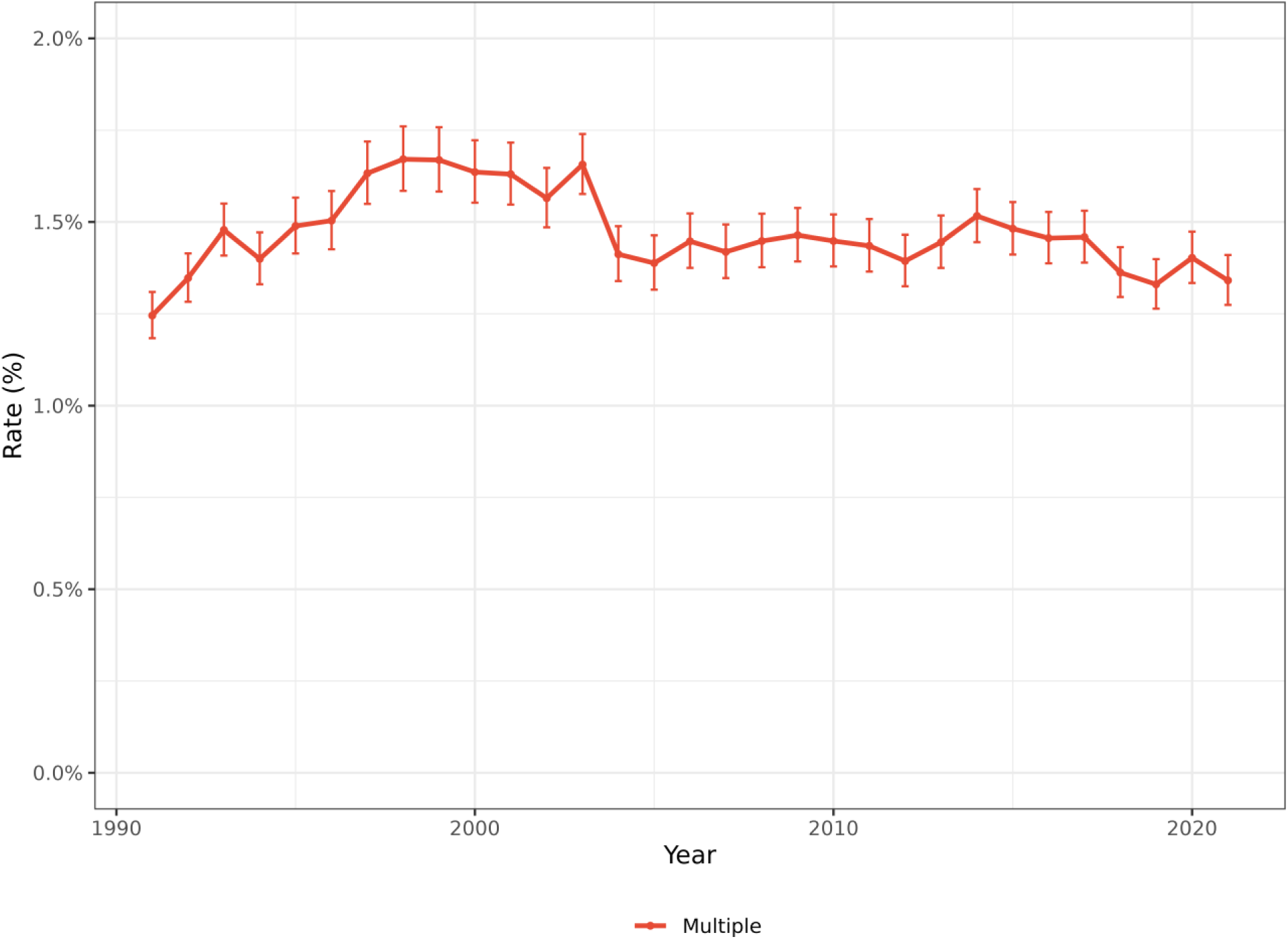
Rate of multiple pregnancies in Sweden from 1991-2021. Error bars indicate 95% CI.

**Supplementary Figure 5.**
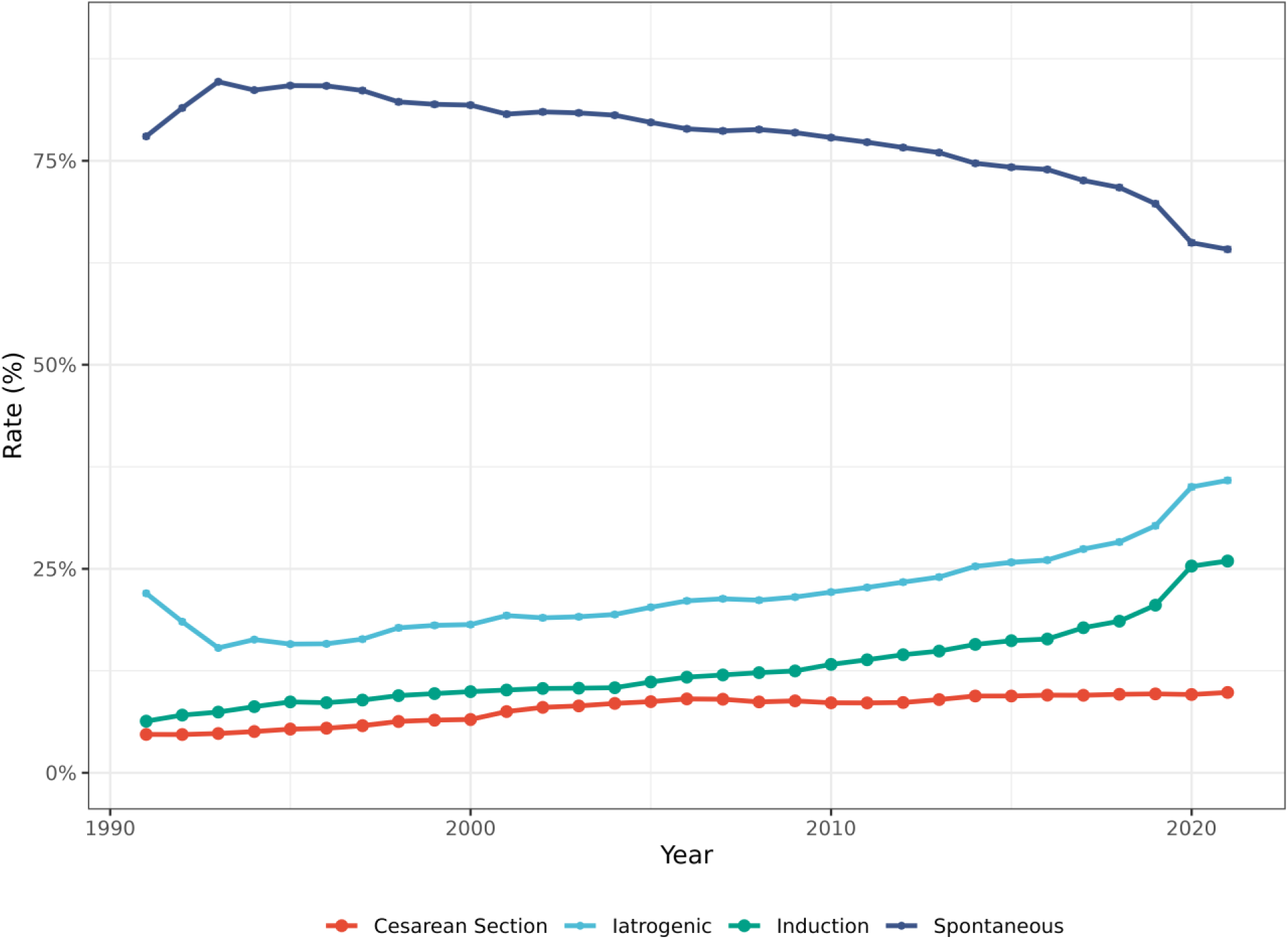
Rates of labour mode in Sweden between 1991-2021. Error bars indicate 95% CI.

**Supplementary Figure 6.**
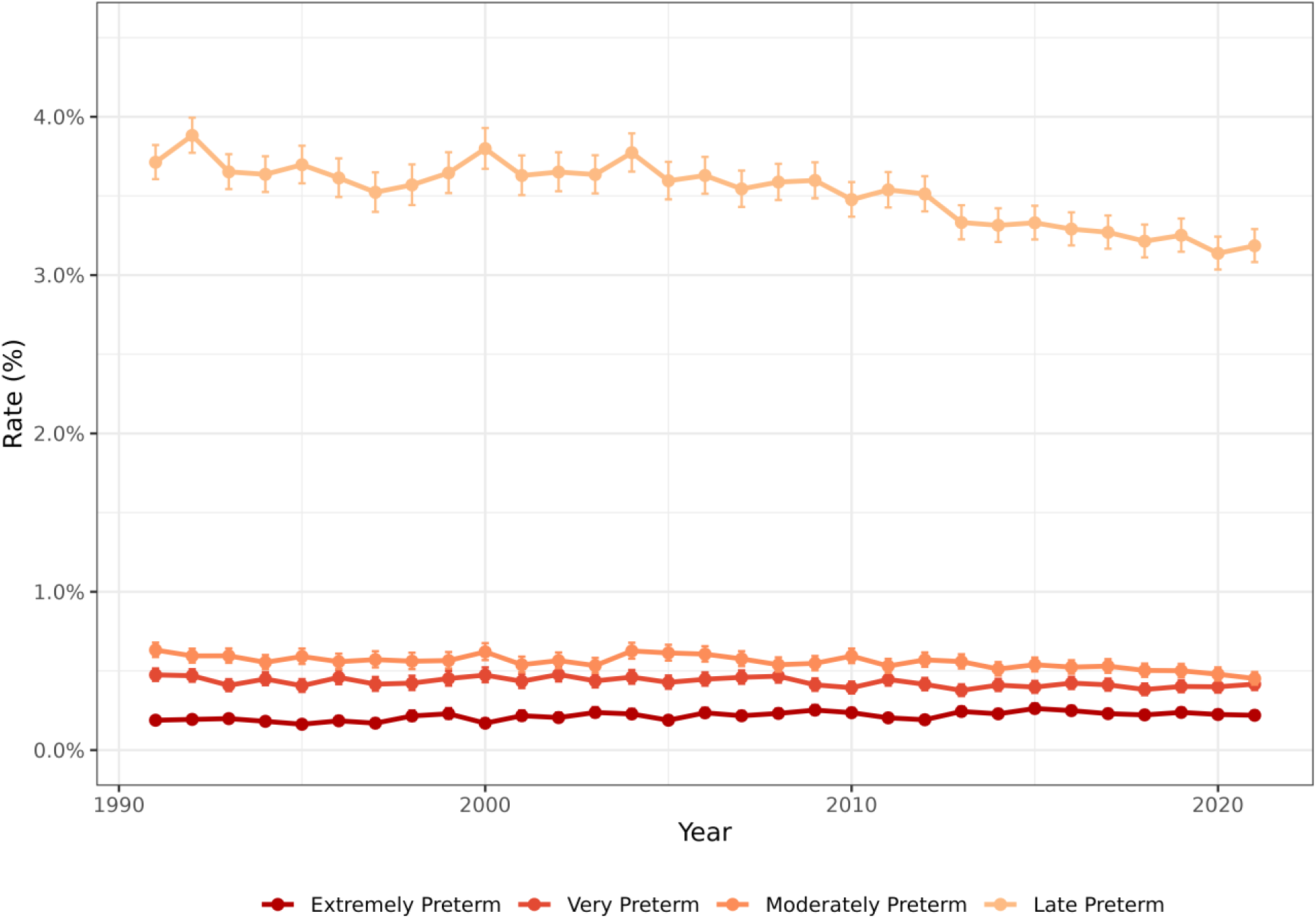
Rates of extremely preterm (<28 weeks, <196 days), very preterm (28-31 weeks, 196 - 224 days), moderately preterm (32 - 33 weeks, 224 - 238 days), and late preterm (34 - 36 weeks, 238 - 259 days) singleton pregnancies in Sweden between 1991-2021. Rates of preterm birth were unadjusted by the number of preterm births over the total number of births in that year. Error bars indicate 95% CI.

**Supplementary Figure 7.**
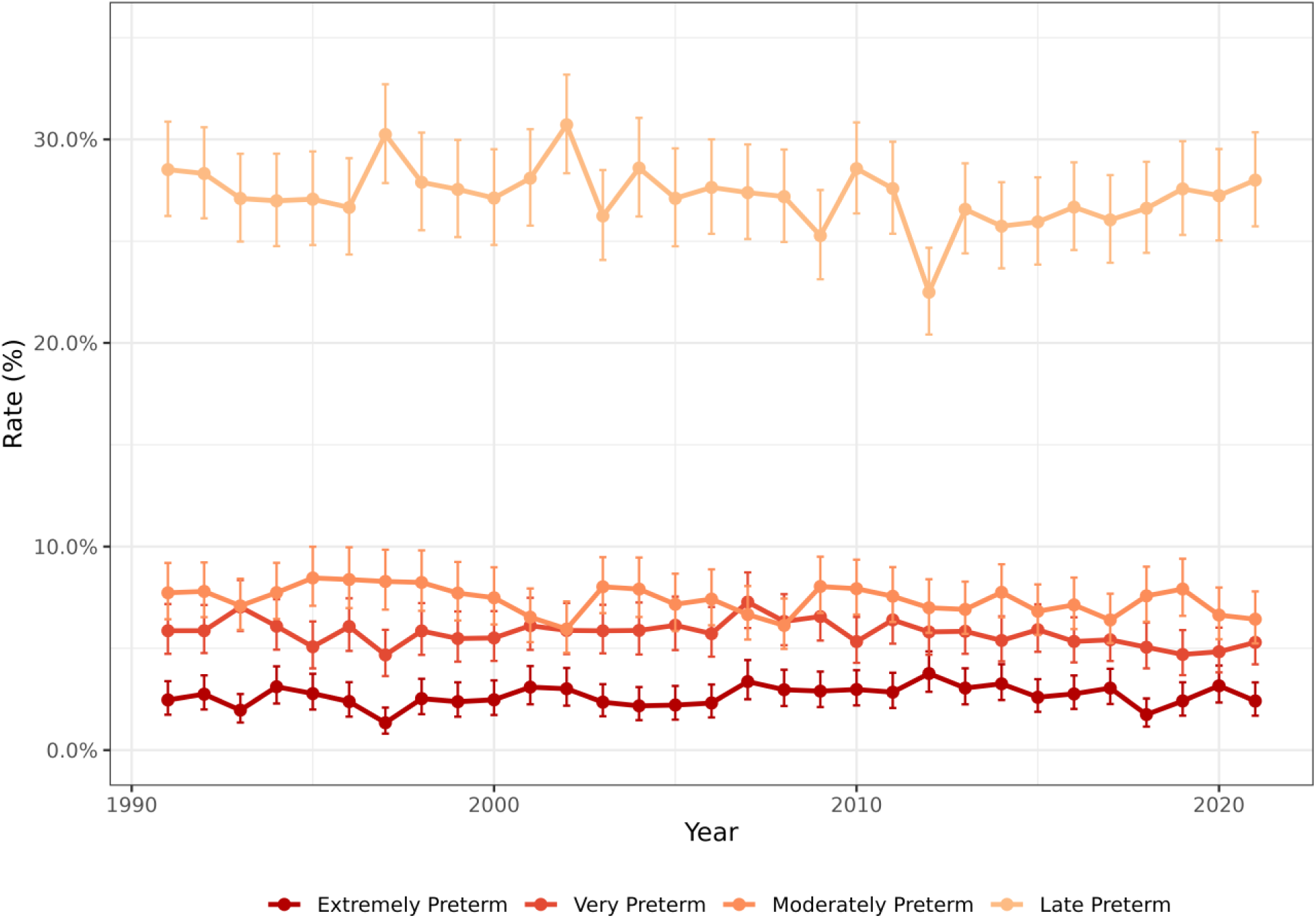
Rates of extremely preterm (<28 weeks, <196 days), very preterm (28-31 weeks, 196 - 224 days), moderately preterm (32 - 33 weeks, 224 - 238 days), and late preterm (34 - 36 weeks, 238 - 259 days) in multiple pregnancies in Sweden between 1991-2021. Rates of preterm birth were unadjusted by the number of preterm births over the total number of births in that year. Error bars indicate 95% CI.

**Figure 8.**
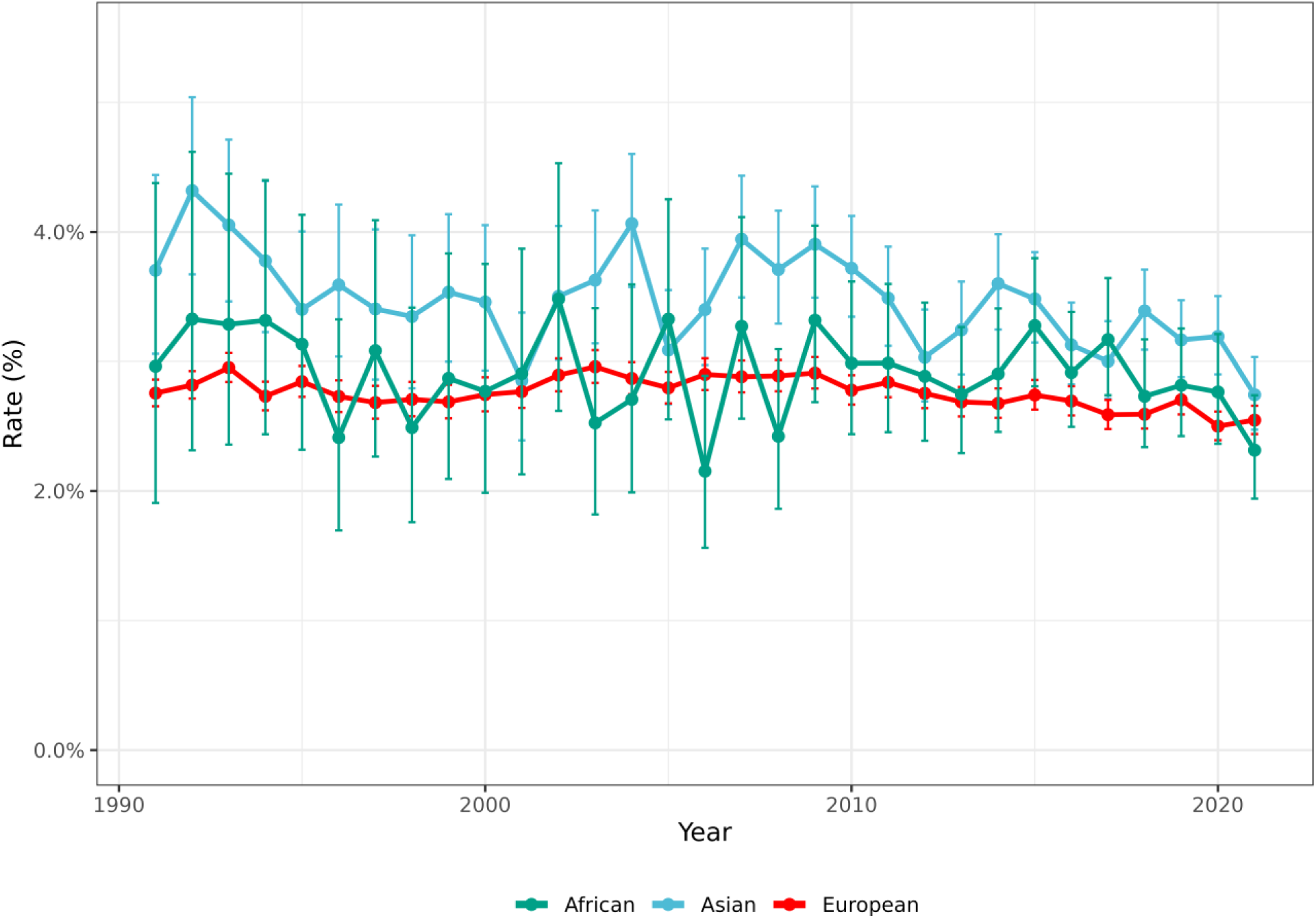
Singleton spontaneous preterm birth rates by last menstrual period in mothers born in Europe, Asia and Africa giving birth in Sweden during the time-period 1991 - 2021. Error bars indicate 95% CI.

## Notes

### Competing Interest Statement

The authors have declared no competing interest.

### Author Declarations

Ethical approval his project was approved by the Swedish Ethical Review Authority (Dnr. 2024-08620-02), allowing the use of pseudonymized individual-level data from the Swedish Medical Birth Register. Because of the nature of the data, informed consent was waived.

